# Slow waning of antibodies following a third dose of BNT162b2 in adults who had previously received two doses of inactivated vaccine

**DOI:** 10.1101/2022.07.18.22277741

**Authors:** Benjamin J. Cowling, Samuel M. S. Cheng, Mario Martín-Sánchez, Niki Y. M. Au, Karl C. K. Chan, John K. C. Li, Leo L. H. Luk, Leo C. H. Tsang, Dennis K. M. Ip, Leo L. M. Poon, Gabriel M. Leung, J. S. Malik Peiris, Nancy H. L. Leung

## Abstract

**Introduction:** Third doses of COVID-19 vaccination provide an important boost to immunity, reducing the risk of symptomatic infection and the risk of severe disease. Third doses have been particularly important for improving protection against variants. However, waning of clinical protection particularly against Omicron has been noted after receipt of third doses.

**Methods:** We administered BNT162b2 as a third dose to adults aged ≥30 years who had previously received two doses of inactivated vaccination. We collected blood before the third dose and again after one month and six months, and tested sera using a spike receptor binding domain IgG enzyme-linked immunosorbent assay, a surrogate virus neutralization test, and live virus plaque reduction neutralization assay against ancestral virus and Omicron BA.2.

**Results:** We administered BNT162b2 as a third dose to 314 adults. We found robust antibody responses to the ancestral strain at six months after receipt of BNT162b2. Antibody responses to Omicron BA.2 were weaker after the third dose and had declined to a low level by six months. From a small number of participants we observed that natural infection or a fourth dose of vaccination generated similar antibody levels against ancestral virus, but infection generated higher antibody level against Omicron BA.2 than vaccination, suggesting a potential advantage in the breadth of antibody response from hybrid immunity.

**Conclusions:** While antibody levels against the ancestral strain remained robust at six months after the third dose, antibody levels against Omicron BA.2 had fallen to low levels suggesting the potential benefits of a fourth dose.

## INTRODUCTION

Third doses of COVID-19 vaccination provide an important boost to immunity, reducing the risk of symptomatic infection [1, 2] and the risk of severe disease [2, 3]. Third doses have been particularly important for improving protection against variants. However, waning clinical protection particularly against Omicron was noted after receipt of third doses [4, 5], with fourth doses then providing additional protection [6, 7].

In recipients of two initial doses of inactivated COVID-19 vaccines, we and others have shown that a third “booster” dose of BNT162b2 (BioNTech/Pfizer/Fosun Pharma) confers a very strong antibody response both against the ancestral strain and the Omicron variant [8-10]. Here, we explore the persistence of antibody titers up to 6 months after a third dose of BNT162b2 in this regimen.

## METHODS

### Study design

We conducted an open-label single-arm trial to measure the antibody responses to a third dose of BNT162b2 in adults ≥30 years old who previously received two doses of an inactivated COVID-19 vaccine with the second inactivated vaccine dose at least 90 days prior to enrolment [8]. The BNT162b2 vaccine and the inactivated vaccine CoronaVac (Sinovac) were both approved for use in Hong Kong in early 2021, while an alternative inactivated vaccine BIBP (Sinopharm) has been available since early 2021 in mainland China and some other countries. We enrolled participants from October through December 2021. Participants were not eligible if they had received any other COVID-19 vaccination apart from the two doses of inactivated vaccination, if they had a history of laboratory-confirmed COVID-19 infection, if they met a contraindication for BNT162b2, were receiving immuno-modulatory medications, or were females who were pregnant or intending to become pregnant in the coming 3 months [8].

Each participant provided a serum sample at Day 0 prior to receipt of BNT162b2, and then further serum samples on Day 28 (±7 days) and Day 182 (±30 days), with a final sample planned on Day 365. Participants were provided with a gift voucher of HK$100 (US$13) at the blood draws on Days 28, 182 and 365. We collected information at baseline on demographics, health status including vaccinations received, and self-reported COVID-19 infection history. We updated this information at the Day 182 visit including information on any infections that had occurred between Day 28 and Day 182.

### Ethical approval

All participants provided written informed consent. The study was approved by the Institutional Review Board of the University of Hong Kong. The study is registered on Clinicaltrials.gov (NCT05057182).

### Laboratory methods

We used a SARS-CoV-2 Spike receptor binding domain IgG enzyme-linked immunosorbent assay (ELISA) for the ancestral strain as previously described [11]. 96-well ELISA plates (Nunc MaxiSorp, Thermo Fisher Scientific) were coated overnight with 100 ng/well of the purified recombinant RBD protein in PBS buffer. The plates were then blocked by 100 μl of Chonblock blocking buffer (Chondrex Inc, Redmond, US) per well, and were incubated at room temperature for 2 hours. Each serum sample was tested at a dilution of 1:100 in Chonblock blocking buffer in duplicate. They were added and were incubated for 2 hours at 37°C. After extensive washing with PBS containing 0.1% Tween 20, horseradish peroxidase (HRP)-conjugated goat anti-human IgG (1:5,000, GE Healthcare) was added and incubated for 1 hour at 37°C. The ELISA plates were then washed again with PBS containing 0.1% Tween 20. Subsequently, 100 μL of HRP substrate (Ncm TMB One; New Cell and Molecular Biotech Co. Ltd, Suzhou, China) was added into each well. After 15 minutes incubation, the reaction was stopped by adding 50 μL of 2□M H_2_SO_4_ solution and analyzed on a microplate reader at 450 nm wavelength. Optical density above 0.5 was counted as positive.

SARS-CoV-2 surrogate virus neutralization test (sVNT) kits (Cat. No.: L00847-A) were ordered from GenScript, Inc., NJ, USA. The tests were performed according to the manufacturer’s standard protocol. 10 times dilution were performed for samples, positive and negative controls. They were then mixed with equal volume of horseradish peroxidase (HRP) conjugated SARS-CoV-2 spike receptor binding domain (RBD) (6 ng). The mixture was incubated at 37°C for 30 min. After incubation, 100ul of the mixture was added to corresponding wells of the capture plate coated with ACE-2 receptor. The plate was sealed and incubated at 37°C for 30 min followed removing mixtures and washing with 1X wash solution four times. Residual liquid was emptied by tapping dry. 100ul of TMB solution was added to each well, the plate was wrapped with aluminium foil and incubated in the dark at room temperature for 15 minutes. The reaction was quenched by adding 50ul of stop solution. The absorbance was read at 450nm (OD_450_) in an ELISA microplate reader. Percentage inhibition was calculated by (1-OD_450_ value of sample/OD_450_ value of negative control) multiplied by 100%.

The Plaque Reduction Neutralization Test (PRNT) was performed in duplicate using 24-well tissue culture plates (TPP Techno Plastic Products AG, Trasadingen, Switzerland) in a biosafety level 3 facility using Vero E6 TMRESS2 cells [12] for the ancestral strain and Omicron BA.2 as previously described [13]. All sera were heat-inactivated at 56°C for 30 min before testing. Serial two-fold dilutions from 1:10 to 1:320 of each serum sample were incubated with 30–40 plaque-forming units of virus for 1 hour at 37°C and the mixture was added onto pre-formed cell monolayers. The culture plate was incubated for 1 hour at 37°C in a 5% CO_2_ incubator. The virus-antibody inoculum was then discarded, and the cell monolayer was overlaid with 1% agarose in cell culture medium. The plates were fixed and stained after 3 days incubation. Antibody titres were defined as the reciprocal of the highest serum dilution that resulted in ≥50% reduction in the number of virus plaques (PRNT_50_). The average plaque numbers were calculated from the duplicates. Virus back titrations, positive and negative control sera were included in every experiment.

### Statistical analysis

We analyzed data on antibody titers measured by the assays listed above at Day 0, Day 28 and Day 182. We determined whether participants reported a laboratory-confirmed infection between Day 28 and Day 182, or had chosen to receive a fourth dose of a COVID-19 vaccine during the same period, and classified accordingly for analysis. We estimated group means for the ELISA and surrogate neutralizing percentages, and geometric mean titers for the live virus neutralization titers. We estimated the rate of waning in neutralizing antibody titers for those who were not infected and did not receive a fourth dose assuming an exponential rate of decline (constant decline on the logarithmic scale). Statistical analyses were conducted using R version 3.6.2 (R Foundation for Statistical Computing, Vienna, Austria).

## RESULTS

We administered BNT162b2 as a third dose of COVID-19 vaccination to 314 participants between 18 October and 28 December 2021. We collected Day 28 samples from 312 (99%) of these participants, and we collected Day 182 samples from 284 (90%) participants between 20 April and 1 June 2022. The median time between receipt of BNT162b2 and collection of the Day 182 sample was 181 days (range 154, 210 days).

Further analyses focus on the 284 participants who provided a Day 182 sample. Among these 284 participants, 279 (98%) had received two initial doses of CoronaVac and the remainder received two doses of BIBP. The median delay between the second dose of inactivated vaccination and the third dose of BNT162b2 administered in our study was 205 days (range 94, 267 days). The median age was 53 years, 29% of participants were ≥60 years of age, 38% were female, and 93 (32%) had a chronic medical condition.

Among the 284 participants, 42 (15%) reported a COVID-19 infection between receipt of the third dose and collection of the Day 182 sample, and 21 (7.0%) reported receipt of a fourth dose prior to collection of the Day 182 sample including one participant who was infected as well as then receiving a fourth dose. The median delay from infection to collection of the Day 182 sample was 55 days (range 10, 85 days). The median delay from the fourth dose to collection of the Day 182 sample was 19 days (range 7, 26) and 19 participants received BNT162b2 as a fourth dose while two received CoronaVac.

The third dose of BNT162b2 led to substantial increases in ELISA (Figure 1A) and surrogate virus neutralization levels (Figure 1B) at Day 28, which waned somewhat by Day 182 but still remained substantially higher than the levels at Day 0. ELISA values at Day 182 were statistically significantly higher in the small number of participants who were infected (p<0.001, t-test) or received a fourth dose (p=0.005, t-test) prior to Day 182. The sVNT responses were very high at Day 182 in all groups, but also statistically significantly higher in the small number of participants who were infected (p=0.002, t-test) or received a fourth dose (p=0.032, t-test) prior to Day 182. There was no statistically significant difference in ELISA or sVNT levels by age at Day 182.

**Figure 1.**
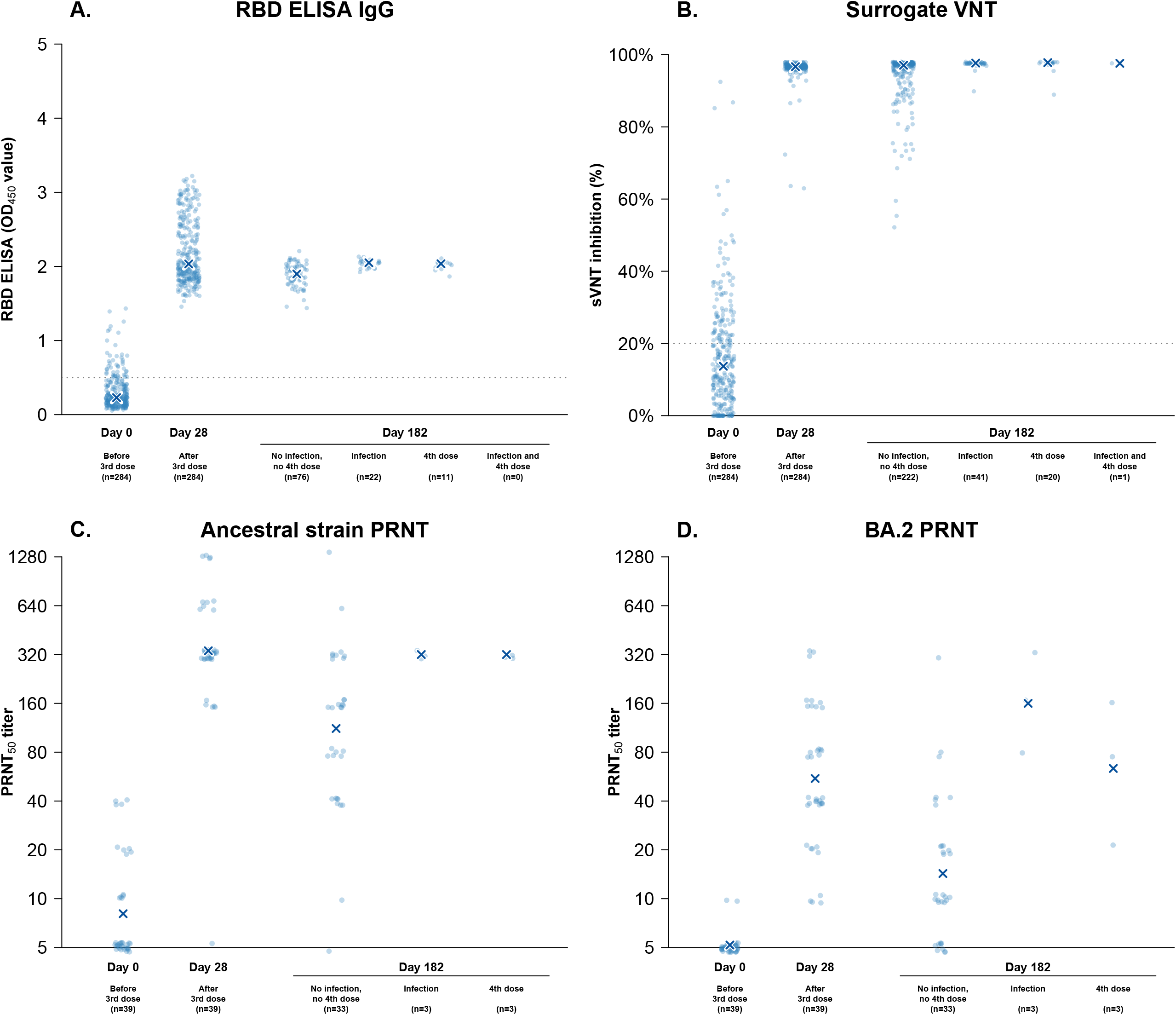
Antibody titers measured prior to receipt of a third dose of BNT162b2 (Day 0), and at Day 28 and Day 182 following that dose, using four assays. Samples collected at Day 182 were stratified by whether the participant had been infected or received a fourth dose between Day 28 and Day 182. Panel A: antibody titers measured by an ELISA assay for serum IgG against the receptor binding domain (RBD) of the spike protein of the ancestral strain, with X indicating the median level. Panel B: Responses to a surrogate virus neutralization test (sVNT) against the ancestral strain, with X indicating the median level. Panel C: Live virus plaque reduction neutralization test (PRNT) against ancestral strain with endpoints at 50% (PRNT_50_) with X indicating the geometric mean titer in each group. Panel D: Live virus PRNT_50_ against the Omicron BA.2 subvariant, with X indicating the geometric mean titer in each group. In panels C and D, antibody titers measured at <10 are plotted at 5 on the y-axis.

We measured PRNT_50_ titers against the ancestral strain (Figure 1C) and Omicron BA.2 (Figure 1D) in a subset of 39 participants at Day 0, 28 and 182. In the statistical comparisons within this subset that follow we exclude (because of the small sample sizes) from Day 182 calculations the three infected participants and the two who received a fourth dose, although they are included in Figure 1 for completeness. At Day 28 and Day 182 the geometric mean PRNT_50_ titers against the ancestral strain were 338 and 112, respectively. The corresponding geometric mean PRNT_50_ titers against BA.2 were 55 and 14, respectively. There was no significant difference by age in PRNT_50_ titers against the ancestral strain or BA.2 at Day 182. Assuming an exponential rate of waning from Day 28 to Day 182, we estimated that antibody titers would drop by half in 96 days for the ancestral strain and 79 days for BA.2.

Among the 42 infections, 30 (71%) occurred during the month of March 2022, with nine in February and three in April, when BA.2 predominated in the community. We did not find any statistically significant effects on the risk of laboratory-confirmed infection of the timing of third dose, age, sex, chronic conditions, or antibody measures at Day 28 (all p>0.05, proportional hazards regression).

## DISCUSSION

We show durable antibody responses to the ancestral strain six months after the third dose of BNT162b2 (Figure 1), consistent with other studies that show a strong and sustained antibody response to a third dose of BNT162b2 after two doses of BNT162b2 [14, 15] or after two doses of inactivated vaccination [16]. Antibody titers measured by sVNT against the ancestral strain were higher at 97% six months after the third dose of BNT162b2 (and two earlier doses of CoronaVac) than six months after either two doses of BNT162b2 or two doses of CoronaVac, when sVNT inhibition had fallen to 80% and 20% respectively in another study [17]. However, neutralizing titers to Omicron BA.2 only reached a moderate geometric mean titer of 55 after the third dose, above a threshold thought to provide some degree of protection against infection in this assay [13] but titers had fallen to below a geometric mean of 14 within six months, likely below the protective threshold (Figure 1D).

There is some evidence from observational studies that third doses can protect against symptomatic Omicron BA.2 infection [1, 2]. In a study of the effectiveness of two and three doses of COVID-19 vaccines in Hong Kong we found evidence suggestive of a moderate level of protection against mild infection [18]. The immune mechanisms contributing to that protection are not fully elucidated but neutralizing antibodies are likely to play a major role, while cellular immunity may also contribute [19]. Fourth doses did improve antibody levels against the ancestral strain (Figure 1) and likely also against BA.2. From a small number of participants we observed that natural infection or (fourth dose) vaccination after third dose BNT162b2 vaccination generated similar antibody levels against ancestral virus, but infection generated higher antibody level against Omicron BA.2 than vaccination, suggesting a potential advantage in the breadth of antibody response from hybrid immunity [20]. Further studies are needed to confirm this finding and to determine the optimal timing of fourth doses under different types of prior immunity.

Our study had a number of limitations. A large wave of Omicron BA.2 occurred in Hong Kong in February-April 2022 with more than 1 million confirmed cases (14% of the population) and 9000 deaths (1.2 deaths per 1000 persons) [21]. Many infections likely were undocumented. While 15% of our cohort reported an infection, including some infections that may not have been documented in the official case count, some other participants may have had an unrecognized infection, biasing upwards the antibody titers at Day 182.

In conclusion, a third dose of BNT162b2 provided a strong and durable immune response in adults who had previously received two doses of inactivated COVID-19 vaccine. Further research is needed on the value of immunogenicity data (including cellular immunity measures as well as antibody levels) to predict the clinical effectiveness of booster doses against symptomatic disease and severe disease with Omicron subvariants.

## Data Availability

All data produced in the present study are available upon reasonable request to the authors

## ACKNOWLEDGEMENTS

We gratefully acknowledge colleagues including Zacary Chai, Sara Chaothai, Kelvin Kwan, Yvonne Ng, Teresa So and Eileen Yu for technical support in preparing and conducting this study; Anson Ho for setting up the database; Julie Au and Lilly Wang for administrative support; Hetti Cheung, Victoria Wong, Bobo Yeung at HKU Health System; Cindy Man and other colleagues at the HKU Community Vaccination Centres at Gleneagles Hospital; and all the study participants for facilitating the study.

## AUTHOR CONTRIBUTIONS

All authors meet the ICMJE criteria for authorship. Each author’s contributions to the paper are listed below according to the CRediT model:

Conceptualization: BJC, GML, NHLL

Methodology: BJC, SMSC

Formal analysis: BJC, MM-S

Investigation: NYMA, KCKC, JKCL, LLHL

Funding acquisition: BJC

Project administration: BJC, SMSC, JSMP, NHLL

Supervision: BJC, SMSC, DKMI, LLMP, GML, JSMP, NHLL

Writing – original draft: BJC

Writing – review & editing: BJC, SMSC, MM-S, NYMA, KCKC, JKCL, LLHL, DKMI, LLMP, GML, JSMP, NHLL.

## REFERENCES

1. Andrews N, Stowe J, Kirsebom F, et al. Covid-19 Vaccine Effectiveness against the Omicron (B.1.1.529) Variant. N Engl J Med 2022; 386:1532–46.

2. Chemaitelly H, Ayoub HH, AlMukdad S, et al. Duration of mRNA vaccine protection against SARS-CoV-2 Omicron BA.1 and BA.2 subvariants in Qatar. Nat Commun 2022; 13:3082.

3. Barda N, Dagan N, Cohen C, et al. Effectiveness of a third dose of the BNT162b2 mRNA COVID-19 vaccine for preventing severe outcomes in Israel: an observational study. Lancet 2021; 398:2093–100.

4. Ferdinands JM, Rao S, Dixon BE, et al. Waning 2-Dose and 3-Dose Effectiveness of mRNA Vaccines Against COVID-19-Associated Emergency Department and Urgent Care Encounters and Hospitalizations Among Adults During Periods of Delta and Omicron Variant Predominance - VISION Network, 10 States, August 2021-January 2022. MMWR Morb Mortal Wkly Rep 2022; 71:255–63.

5. Patalon T, Saciuk Y, Peretz A, et al. Waning effectiveness of the third dose of the BNT162b2 mRNA COVID-19 vaccine. Nat Commun 2022; 13:3203.

6. Bar-On YM, Goldberg Y, Mandel M, et al. Protection by a Fourth Dose of BNT162b2 against Omicron in Israel. N Engl J Med 2022; 386:1712–20.

7. Grewal R, Kitchen SA, Nguyen L, et al. Effectiveness of a fourth dose of covid-19 mRNA vaccine against the omicron variant among long term care residents in Ontario, Canada: test negative design study. BMJ 2022; 378:e071502.

8. Leung NHL, Cheng SMS, Martin-Sanchez M, et al. Immunogenicity of a third dose of BNT162b2 to ancestral SARS-CoV-2 & Omicron variant in adults who received two doses of inactivated vaccine. Clin Infect Dis 2022.

9. Cheng SMS, Mok CKP, Leung YWY, et al. Neutralizing antibodies against the SARS-CoV-2 Omicron variant BA.1 following homologous and heterologous CoronaVac or BNT162b2 vaccination. Nat Med 2022; 28:486–9.

10. Campos GRF, Almeida NBF, Filgueiras PS, et al. Booster dose of BNT162b2 after two doses of CoronaVac improves neutralization of SARS-CoV-2 Omicron variant. Commun Med (Lond) 2022; 2:76.

11. Perera RAPM, Mok CKP, Tsang OTY, et al. Serological assays for SARS-CoV-2. Eurosurveillance 2020 (in press).

12. Matsuyama S, Nao N, Shirato K, et al. Enhanced isolation of SARS-CoV-2 by TMPRSS2-expressing cells. Proc Natl Acad Sci U S A 2020; 117:7001–3.

13. Lau EH, Hui DS, Tsang OT, et al. Long-term persistence of SARS-CoV-2 neutralizing antibody responses after infection and estimates of the duration of protection. EClinicalMedicine 2021; 41:101174.

14. Gilboa M, Regev-Yochay G, Mandelboim M, et al. Durability of the immune response to a third BNT162b2 dose; five months follow-up. medRxiv 2022:doi: 10.1101/2022.05.03.22274592.

15. Eliakim-Raz N, Stemmer A, Leibovici-Weisman Y, et al. Durability of response to the third dose of the SARS-CoV-2 BNT162b2 vaccine in adults aged 60 years and older: Three-month follow-up. medRxiv 2022:doi: 10.1101/2021.12.25.21268336.

16. Silva ARD, Jr., Villas-Boas LS, Paula AV, et al. Neutralizing antibodies against the SARS-CoV-2 Omicron variant following two CoronaVac vaccinations and a Pfizer/BioNTech mRNA vaccine booster. Rev Inst Med Trop Sao Paulo 2022; 64:e43.

17. Cowling BJ, Wong IOL, Shiu EYC, et al. Strength and durability of antibody responses to BNT162b2 and CoronaVac. Vaccine 2022.

18. McMenamin ME, Nealon J, Lin Y, et al. Vaccine effectiveness of two and three doses of BNT162b2 and CoronaVac against COVID-19 in Hong Kong. Lancet Infect Dis 2022.

19. Kent SJ, Khoury DS, Reynaldi A, et al. Disentangling the relative importance of T cell responses in COVID-19: leading actors or supporting cast? Nat Rev Immunol 2022; 22:387–97.

20. Epsi NJ, Richard SA, Lindholm DA, et al. Understanding ‘hybrid immunity’: comparison and predictors of humoral immune responses to SARS-CoV-2 infection and COVID-19 vaccines. Clin Infect Dis 2022.

21. Mefsin Y, Chen D, Bond HS, et al. Epidemiology of infections with SARS-CoV-2 Omicron BA.2 variant in Hong Kong, January-March 2022. medRxiv 2022:2022.04.07.22273595.

